# The indirect effect of mRNA-based Covid-19 vaccination on unvaccinated household members

**DOI:** 10.1101/2021.05.27.21257896

**Authors:** Jussipekka Salo, Milla Hägg, Mika Kortelainen, Tuija Leino, Tanja Saxell, Markku Siikanen, Lauri Sääksvuori

**Affiliations:** Faculty of Social Sciences, University of Helsinki, VATT, Aalto University and Helsinki GSE, Arkadiankatu 7, 00101 Helsinki, Finland; Faculty of Social Sciences, University of Helsinki, VATT, Aalto University and Helsinki GSE, Arkadiankatu 7, P.O. Box 1279, 00101 Helsinki, Finland; Department of Economics, Turku School of Economics, InFLAMES Research Flagship Center, University of Turku, VATT and Helsinki GSE, Rehtorinpellonkatu 3, 20500 Turku, Finland; Finnish Institute for Health and Welfare, Infectious Disease Control and Vaccinations Unit, P.O. Box 30, FI-00271 Helsinki, Finland; VATT and Helsinki GSE, Arkadiankatu 7 P.O. Box 1279, 00101 Helsinki, Finland; VATT and Helsinki GSE, Arkadiankatu 7, P.O. Box 1279, 00101 Helsinki, Finland; Finnish Institute for Health and Welfare, Centre for Health and Social Economics and University of Turku, INVEST Research Flagship Center, Assistentinkatu 7, 20014 University of Turku, Finland

## Abstract

Mass vaccination is effective in reducing SARS-CoV-2 infections among vaccinated individuals. However, it remains unclear how effectively Covid-19 vaccines prevent people from spreading the virus to their close contacts in real-world circumstances. Here, using nationwide administrative datasets on SARS-CoV-2 infections, vaccination records, demographics, and unique household IDs, we provide evidence about the direct and indirect effectiveness of Covid-19 vaccines in reducing infections among vaccinated individuals and their unvaccinated household members. Our estimates for adults imply indirect effectiveness of 8.7% (95% CI: -28.9 to 35.4) two weeks and 42.9% (95% CI: 22.3 to 58.1) 10 weeks after the first dose. We find that the indirect effect of Covid-19 vaccines is smaller and less precise for unvaccinated children aged between 3 to 18 years than for adults. These results provide household-level evidence that vaccines do not only protect vaccinated individuals but provide indirect protection to unvaccinated individuals in a real-world setting.

---

Governments around the world are hoping to lift Covid-19 restrictions as vaccination coverage increases. However, with the limited supply of vaccines and the emergence of new virus variants, decision-makers in public health are facing a constant need to update their vaccination and contact restriction strategies. There is currently no consensus as to whether to include all children in mass vaccination programs and much uncertainty about the prospect of achieving herd immunity through vaccination.

Covid-19 vaccines have been shown to be effective in preventing symptomatic and asymptomatic disease among vaccinated adults both in clinical trials and in nationwide mass vaccination settings.^1–4^ Furthermore, there is evidence that vaccinated individuals infected with SARS-CoV-2 have lower viral loads than unvaccinated infected individuals.^5^ However, it remains unclear how effectively Covid-19 vaccines prevent people from becoming infected and spreading the virus to their close contacts, most notably to children and other household members, in real-world circumstances.

Here, we provide evidence about the direct and indirect effectiveness of Covid-19 vaccines in reducing infections among vaccinated individuals and their unvaccinated household members. Our analysis exploits the rollout of the mass vaccine program in a large cohort of healthcare workers in Finland, allowing us to estimate the indirect effects of vaccines in a large sample of household members with discordant vaccination status.

We used national databases that record all polymerase chain reaction (PCR)-confirmed SARS-CoV-2 infections and mRNA-based (BNT162b2 by Pfizer-BioNTech or mRNA-1273 by Moderna) vaccine doses administered in Finland since the beginning of the epidemic. These data were merged with administrative full-population datasets that include information on each person’s occupation and unique identifiers for spouses and children living in the same household. To estimate the direct and indirect effectiveness of mRNA-based vaccines, we compared the cumulative incidence of SARS-CoV-2 infections between vaccinated and unvaccinated healthcare workers as well as between their unvaccinated spouses and children living in the same household (see Online content, Tables A2 and A3, for the covariate balance between the treatment and control groups). The estimates were adjusted for the state of the epidemic (calendar time), age and sex.

We report vaccine effectiveness estimates by follow-up week after receiving the first dose. For each unvaccinated healthcare worker and their family member in the control group, we randomly assigned the beginning of a follow-up period during the observation period (see Online content, Section D, for details). Using this novel approach, we ensure that the estimates are not biased by the changing nature of the epidemic during the follow-up period. Our results remain qualitatively unaltered if we follow all individuals in the control group from the beginning of the mass vaccination program (Online content, Figures A6 and A7). Moreover, to make individuals in the treatment and control groups comparable in terms of their SARS-CoV-2 history, we focused only on persons who had not been infected before their follow-up period. Following the current standard practice in Covid-19 vaccine effectiveness reporting, we report vaccine effectiveness as relative risk reduction.^6^ We used a log-binomial model as it allows direct estimation of the risk reduction. We assessed the robustness of our results using different estimation methods (Online content, Section E).

Figure 1 shows our effectiveness estimates for working-age healthcare workers and their unvaccinated spouses between the start of the mass vaccination program (December 27, 2020) and March 24, 2021 in Finland. Our estimates imply a gradual increase in direct vaccine effectiveness over time (Figure 1, Panel A): 26.8% (95% CI: 7.5 to 42.1) two weeks and 69.0% (95% CI: 59.2 to 76.3) 10 weeks after the first dose. These estimates are consistent with the results from clinical trials and previous observational studies assessing first-dose mRNA-vaccine effectiveness. Our results document vaccine effectiveness in a real-world setting, in which it is difficult to separate the longer-term effects of first- and second-dose vaccinations due to selection in vaccine administration (the second dose of the mRNA vaccine is administered only to individuals with no recent infections). We document that over 40 percent of individuals included in our sample received their second dose four weeks after the first dose (Online content, Figure A1).

**Figure 1:**
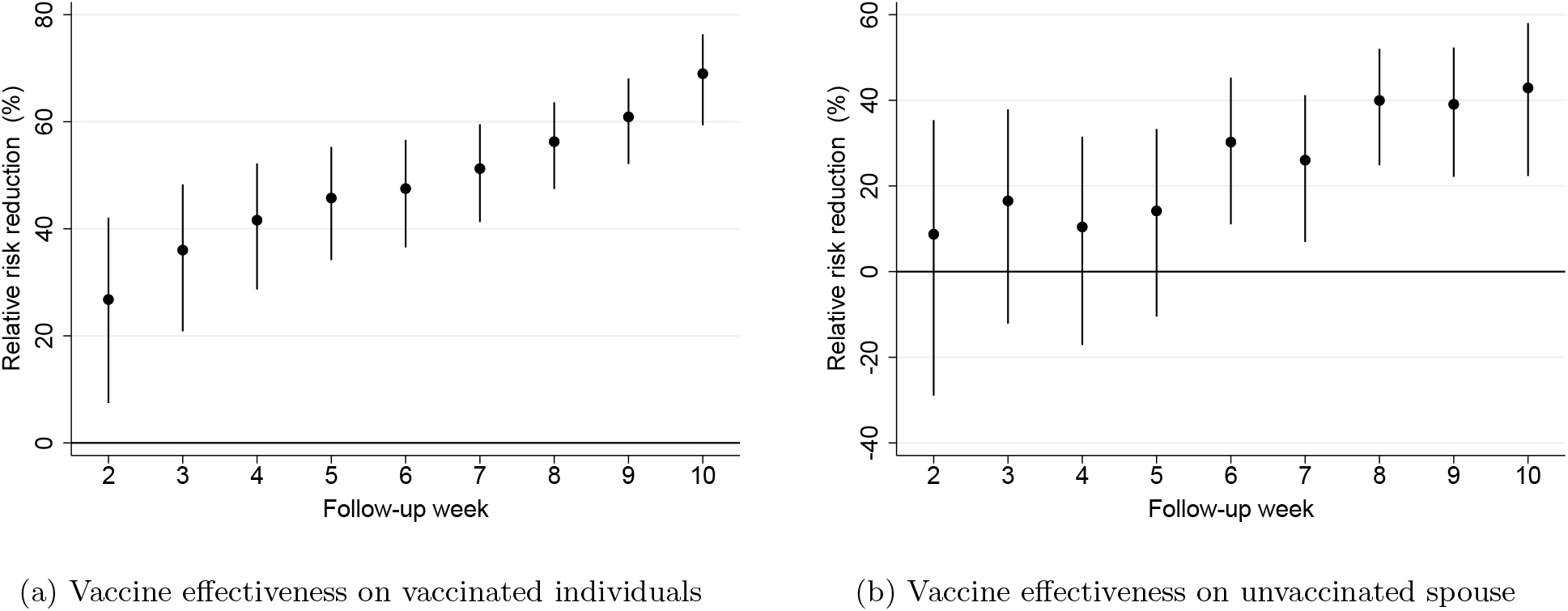
Vaccine effectiveness on vaccinated individuals and unvaccinated spouses living in the same household *Notes:* This figure plots the vaccine effectiveness estimates by week after the first dose of mRNA-based vaccination. The dependent variable is a polymerase chain reaction (PCR)-confirmed SARS-CoV-2 infection as recorded in the Finnish National Infectious Diseases Register. **Panel A** shows our direct effectiveness estimates (relative risk reduction) for vaccinated individuals compared to the control group, which is constructed by randomly assigning a placebo vaccination date for non-vaccinated individuals. **Panel B** shows the effectiveness estimates (relative risk reduction) for the non-vaccinated spouses (including cohabiting partners) of vaccinated individuals who lived in the same household with the vaccinated person as of December 31, 2019. See Online Appendix for details on sample restrictions.The coefficients in both panels are estimated using individual-date data collapsed to individual-week data. To construct the relative risk reduction (RRR) estimates, we first run a log-binomial regression model using a vaccination status indicator variable (vaccinated vs. unvaccinated) by the event week (follow-up week after vaccine administration) and transform the obtained estimates into risk ratio coefficients. We then compute the relative risk reduction using the formula RRR = 1 - risk ratio. The solid black dots show relative risk reduction by week after receiving the first dose of mRNA-based vaccination. The error bars denote the 95% confidence intervals clustered at the individual level using endpoint transformation of the originally estimated confidence intervals.

Our estimates for unvaccinated spouses imply indirect effectiveness of 8.7% (95% CI: -28.9 to 35.4) two weeks and 42.9% (95% CI: 22.3 to 58.1) 10 weeks after the first dose (Figure 1, Panel B). The indirect effectiveness estimates are substantial, but, as expected, are smaller than the direct effect and occur more gradually among unvaccinated spouses than among vaccinated individuals.

Figure 2 shows household-level evidence for the indirect effectiveness of mRNA-based vaccines for preventing SARS-CoV-2 infections among unvaccinated children aged between 3 and 18 years (Panel A) and by age groups: from 3 to 12 years old and from 13 to 18 years old (Panel B). The indirect effectiveness among unvaccinated children aged from 3 to 18 years is estimated to be smaller than among unvaccinated spouses: -1 % (95% CI: -53.9 to 33.7) two weeks and 32.9% (95% CI: 4.1 to 53.0) 10 weeks after the first dose. However, the estimates are more imprecise due to the smaller sample size and the likelihood of infection among children. Figure 2 (Panel B) stacks the estimates for the two age groups from follow-up weeks 2 to 5 into a single estimate as there are too few PCR-confirmed infections among children aged from 3 to 12 years to make reliable inference about the short-term indirect effectiveness of Covid-19 vaccines in this age group. The indirect effectiveness estimates for 3 - 12-year-old children are not statistically significantly different from zero: 12.3% (95% CI: -37.4 to 44.0) 6 weeks and 22.3% (95% CI: -34.4 to 55.2) 10 weeks after the first dose. For 13 - 18-year-old adolescents, we find indirect effects that are consistent with the indirect effects for unvaccinated spouses: 16.7% (95% CI: -17.7 to 41.0) 6 weeks and 38.0% (95% CI: 1.2 to 61.1) 10 weeks after the first dose. However, in this age group, the estimates are imprecise and statistically significant only in the tenth follow-up week.

**Figure 2:**
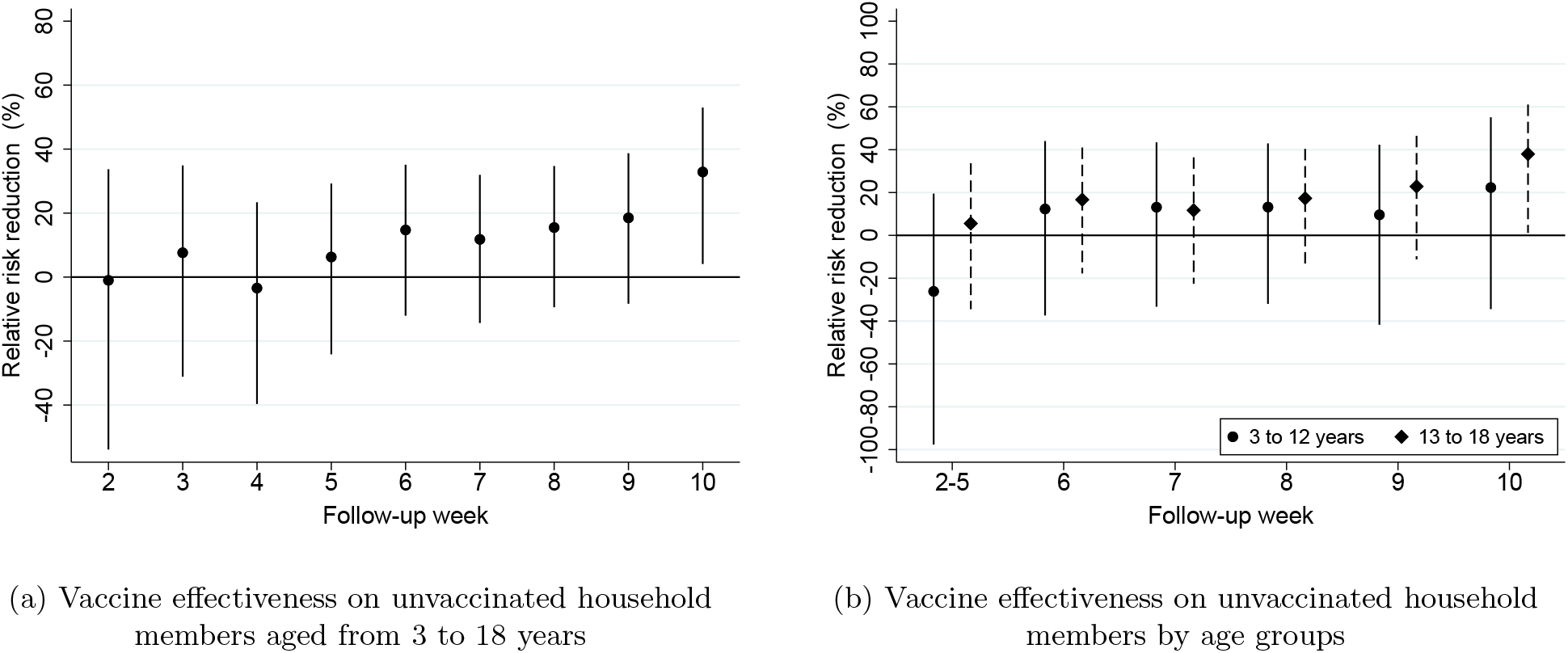
Vaccine effectiveness on unvaccinated children and adolescents *Notes:* This figure plots the vaccine effectiveness estimates by week after the first dose of mRNA-based vaccination. The dependent variable is a polymerase chain reaction (PCR)-confirmed SARS-CoV-2 infection as recorded in the Finnish National Infectious Diseases Register. **Panel A** shows effectiveness estimates (relative risk reduction) for unvaccinated household members aged from 3 to 18 years who lived in the same household with the vaccinated person as of December 31, 2019. **Panel B** shows effectiveness estimates (relative risk reduction) separately for unvaccinated children aged from 3 to 12 years and adolescents aged from 13 to 18 years who lived in the same household with the vaccinated person as of December 31, 2019. Panel B stacks the estimates from follow-up weeks 2 to 5 into a single estimate as there are too few polymerase chain reaction (PCR)-confirmed SARS-CoV-2 infections among children aged from 3 to 12 years to make any inference about the short-term effectiveness of mRNA-based vaccination in this age group. See Online Appendix for details on sample restrictions. The coefficients in both panels are estimated using individual-date data collapsed to individual-week data. To construct the relative risk reduction (RRR) estimates, we first run a log-binomial regression model using a vaccination status indicator variable (vaccinated vs. unvaccinated) by the event week (follow-up week after vaccine administration) and transform the obtained estimates into risk ratio coefficients. We then compute the relative risk reduction using the formula RRR = 1 - risk ratio. The solid black dots show relative risk reduction by week after receiving the first dose of mRNA-based vaccination. The error bars denote the 95% confidence intervals clustered at the household level using endpoint transformation of the originally estimated confidence intervals.

The present study is uniquely suited for evaluating the effect of mRNA-based vaccines on SARS-CoV-2 infections and secondary transmission from vaccinated to unvaccinated individuals. We were able to merge individual-level health records with full-population datasets containing detailed information about vaccinated individuals’ occupations, spouses, and children. Moreover, our research strategy capitalized on the early vaccination of healthcare workers that created a large group of families with discordant vaccination status, critical for the estimation of household spillover effects.

Observational studies on the indirect effectiveness of mRNA-based vaccines have so far paid little attention to children.^7,8^ A notable exception is a community-level study in Israel, which concentrates on the regional effects of vaccinated adults on unvaccinated children’s infections.^9^ By contrast, here we concentrate on evaluating the effect of vaccines on transmission from vaccinated adults to unvaccinated adults and children within the same household.

Methodologically, we estimate indirect vaccine effectiveness by comparing individuals in a treatment versus a control group (defined by the vaccination status of their close contact), mimicking the RCT design. Thus, our research design complements existing observational studies that have relied on comparisons of treated individuals at different periods of time before and after of the vaccine administration to their close contacts.^7,10^

Our study has several important limitations. First, our study is limited by the fact that healthcare workers may differ from the general population, for example, in terms of their SARS-CoV-2 virus exposure. Second, the virus exposure could have differed between vaccinated and unvaccinated healthcare workers as persons directly involved with Covid-19 patients were more likely to be vaccinated. However, our results for the direct vaccine effectiveness after the first dose are very close to the results from randomized controlled clinical trials and prior observational studies, suggesting that the sample selection and the non-random assignment of vaccines to healthcare workers do not introduce substantial bias into our vaccine effectiveness estimates. Third, our study is limited by the fact that we lack data on the potential biological and behavioral mechanisms that could clearly pinpoint the mechanism leading to the observed reduction in confirmed SARS-CoV-2 infections among unvaccinated household members.

Taken together, our results suggest that mRNA-based Covid-19 vaccines do not only prevent SARS-CoV-2 infections among vaccinated individuals but lead to a substantial reduction in infections among unvaccinated adults and children living in the same household. Our results are consistent with the notion that mRNA-based vaccines affect susceptibility in vaccinated individuals and prevent transmission from vaccinated to unvaccinated individuals within households. The observed reduction in household transmission from vaccinated to unvaccinated individuals is expected to curb the current Covid-19 pandemic as household transmission of SARS-CoV-2 is believed to have a significant role in the overall spread of infections in the community.^11–13^ Nevertheless, further studies are required to understand how the indirect effects of Covid-19 vaccines on unvaccinated adults and children support the prospect of herd immunity and to inform questions related to the possible mass vaccination of children.

## Data Availability

This paper uses administrative health care and employment data maintained by the Finnish Institute
for Health and Welfare and Statistics Finland. Health care data is regulated under the Act on the
Secondary Use of Health and Social Data (552/2019) and can be obtained by sending a direct
request to the Finnish Institute for Health and Welfare (https://thl.fi/en). The Finnish Longitudinal Employer-Employee Data (FOLK) can be obtained by sending a direct request to Statistics Finland (https://www.stat.fi). The authors are willing to assist in making data access requests.

## Author contributions

Study design: JS, MK, TS and LS. Data collection: JS, MH and LS. Data analysis: JS and LS. Data interpretation: JS, MH, MK, TL, TS, MS and LS. Writing: JS, MH, MK, TS, MS and LS.

## Acknowledgements

This research was supported by the InFLAMES and INVEST Flagship Programmes of the Academy of Finland.

