## Supplementary material for "The indirect effect of mRNA-based Covid-19 vaccination on unvaccinated household members": Online Appendix

#### Contents

**A Data Sources** 2

**B Healthcare Worker Definition** 2

**C Estimation Sample** 2

**D Estimation Method** 6

**E Supplementary Results** 7

*Salo: Faculty of Social Sciences, University of Helsinki, VATT and Helsinki GSE, Arkadiankatu 7, 00101 Helsinki, Finland.. Hägg: Faculty of Social Sciences, University of Helsinki, VATT and Helsinki GSE, Arkadiankatu 7, P.O. Box 1279, 00101 Helsinki, Finland.. Kortelainen: Department of Economics, Turku School of Economics, InFLAMES Research Flagship Center, University of Turku, VATT and Helsinki GSE, Rehtorinpellonkatu 3, 20500 Turku, Finland.. Leino: Finnish Institute for Health and Welfare, Infectious Disease Control and Vaccinations Unit, P.O. Box 30, FI-00271 Helsinki, Finland.. Saxell: VATT and Helsinki GSE, Arkadiankatu 7 P.O. Box 1279, 00101 Helsinki, Finland.. Siikanen: VATT and Helsinki GSE, Arkadiankatu 7, P.O. Box 1279, 00101 Helsinki, Finland.. Sääksvuori: Finnish Institute for Health and Welfare, Centre for Health and Social Economics and University of Turku, INVEST Research Flagship Center, Assistentinkatu 7, 20014 University of Turku, Finland.. Corresponding author: Lauri Sääksvuori. This research was supported by the InFLAMES and INVEST Flagship Programmes of the Academy of Finland. Dr. Kortelainen declares a grant to his employer, but no personal support or financial relationship, from Pfizer during the conduct of the study. The other authors have no conflicts of interest to declare.

### A Data Sources

The analyses were conducted using multiple population-wide administrative datasets linked at the individual level. All polymerase chain reaction (PCR)-confirmed SARS-CoV-2 infections and their dates before March 24, 2021 were recorded in the Finnish National Infectious Diseases Register. The Finnish National Vaccination Register provided information about the type of vaccine and date of vaccine administration for all Covid-19 vaccines administered before March 18, 2021. We used the Finnish Incomes Register for the year 2020 from the Finnish Tax Authority to identify healthcare workers following Statistics Finland's classification of occupations. These occupations are presented in Section B.

We merged these data with the Statistics Finland FOLK module (full population data) for year 2019 to identify spouses (including cohabiting partners) and children who lived in the same household as the healthcare workers on December 31, 2019. Children born after 2018 cannot be included in the analysis, since we cannot link these children with their parents. Finally, using these linked data, we constructed the outcome variable, the cumulative indicator of SARS-CoV-2 infection, for healthcare workers and their household members used in the statistical analysis of vaccine effectiveness. The outcome variable varies at the individual level for healthcare workers and their spouses and at the household level in the analysis related to children.

### B Healthcare Worker Definition

Our healthcare worker definition was based on the occupation classification of Statistics Finland (TK10). Table A1 shows the occupations which we use to define healthcare workers in our sample. We used 3-digit occupation codes to identify healthcare professions. Some 5-digit occupation codes within these 3-digit codes were excluded, because a significant share of individuals within these occupations may work outside the healthcare sector.

### C Estimation Sample

Our analysis used three distinct estimation samples. The first sample was used in estimating the direct effect of the mRNA-based vaccinations (BNT162b2 by Pfizer-BioNTech or mRNA-1273 by Moderna). This sample consisted of working-age healthcare personnel (age 15 - 74 years). The second sample was used in estimating the indirect vaccine spillover effect on the spouses of the healthcare workers. An individual was included in this sample if their spouse is a healthcare worker and they had not been vaccinated during the sample period. The third sample was used in estimating the indirect vaccine spillover effect on the children

Table A1: Healthcare Worker Occupation Codes

| Occupation code | Included occupations | Excluded occupations (codes) |
|---|---|---|
| 221 | Physicians | |
| 222 | Senior nursing officials, Ward sisters | |
| 322 | Nurses, Midwifes | |
| 226 | Dentists, Audiologists and Speech therapists | Pharmacists (22620), Environmental and occupational health and hygiene professionals (22630), Dieticians and nutritionists (22650), Health professionals not elsewhere classified (22690) |
| 532 | Practical nurses | Pharmaceutical assistants (53293), Equipment maintenance assistants (53292), Massage therapists and practical rehabilitation nurses (53294) |

*Notes:* This table shows the occupations we use to define healthcare workers in our sample. Some occupations are excluded, because a significant share of individuals in these occupations may work outside the healthcare sector. Occupation codes are based on Statistics Finland's classification of occupations (TK10).

Table A2: Descriptive Statistics for Healthcare Workers and Their Spouses

| *Panel A:* | Vaccinated persons (N = 95,138) | Unvaccinated persons (N = 193,000) |
|---|---|---|
| Age - mean | 47.1<br>(13.1) | 43.8<br>(14.5) |
| Women - N | 82,287<br>[86.5%] | 166,898<br>[86.4%] |

| *Panel B:* | Spouses of vaccinated persons (N = 52,766) | Spouses of unvaccinated persons (N = 111,000) |
|---|---|---|
| Age - mean | 48.9<br>(12.4) | 47.0<br>(13.8) |
| Women - N | 5650<br>[10.7%] | 12,956<br>[11.7%] |

*Notes:* This table presents descriptive statistics for the vaccinated and unvaccinated healthcare workers and their spouses. Square brackets report proportions (%) and parentheses show standard deviation.

Table A3: Descriptive Statistics for Oldest Children and by Their Age Group

| *Panel A:* (Age 3 - 18) | Children of vaccinated persons (N = 28,666) | Children of unvaccinated persons (N = 63,987) |
|---|---|---|
| Age - mean | 12.451 | 11.74 |
| | (4.447) | (4.657) |
| Women - N | 14091 | 30912 |
| | [49.2%] | [48.3%] |

| *Panel B:* (Age 3 - 12) | Children of vaccinated persons (N = 12,834) | Children of unvaccinated persons (N = 32,568) |
|---|---|---|
| Age - mean | 8.2 | 7.8 |
| | (2.8) | (2.9) |
| Women - N | 6266 | 15,689 |
| | [48.8%] | [48.2%] |
| *Panel C:* (Age 13 - 18) | Children of vaccinated persons (N = 15,832) | Children of unvaccinated persons (N = 31,419) |
| Age - mean | 15.9 | 15.8 |
| | (1.7) | (1.7) |
| Women - N | 7825 | 15,223 |
| | [49.4%] | [48.5%] |

*Notes:* This table presents the household level descriptive statistics for the healthcare workers' oldest children. Age and sex (Women) variables reported in this table denotes the age and sex of the oldest child in the household. Totals reported in this table denote the number of households Panel A reports statistics for children and adolescents, Panel B for 3 - 12 years old children and Panel C 13 - 18 years old adolescents. Square brackets report proportions (%) and parentheses show standard deviation.

(age 3 - 18) living in the same household as the healthcare worker. Children included in this sample are biological children of vaccinated healthcare workers. An individual was included in this sample if they live in the same household as the healthcare worker and they had not been vaccinated during the sample period.

Tables A2 and A3 document descriptive statistics for the estimation samples and show that 33% of the healthcare workers were vaccinated and 67% were unvaccinated during the observation period. All variables were fairly well balanced between the treatment and control groups. The mean age of the vaccinated healthcare workers was 47 years (SD 13) and for the unvaccinated controls it was 44 years (SD 15). The mean age was 49 years (SD 12) for the treated spouses of vaccinated healthcare workers, and for the controls (the spouses of unvaccinated healthcare workers) it was 47 years (SD 14). It is important to note that descriptive statistics reported in Table A3 are household-level statistics, because the statistical analysis related to children is conducted at this level. If a healthcare worker has many children living in the same household, the table reports the summary statistics of the oldest child. The mean age for the (oldest) children of vaccinated healthcare workers was 12.5 years (SD 4.4) and for the children of unvaccinated healthcare workers the mean age was 11.7 (SD 4.7). As expected, healthcare workers were typically female (Table A2 - Panel A) and their spouses male (Table A2 - Panel B) and the sex of the children is evenly balanced (Table A3 - Panel A). The sample sizes for the spouses and children are smaller than the sample for the healthcare workers. The reasons for this are that some healthcare workers might not have a spouse, the spouse or the children are vaccinated or the healthcare worker might not have any children.

Moreover, Table A4 (Panel A) shows that 31% of vaccinated healthcare workers were nurses and midwives (occupation code 322), and 10% were physicians (occupation code 221). Table A4 (Panel B) shows that 4% of unvaccinated healthcare workers were physicians and 21% were nurses and midwives. The analysis sample mainly consists of practical nurses (occupation code 532) and nurses and midwifes (occupation code 322) in the groups of vaccinated and unvaccinated healthcare workers.

Table A4: Occupations of Vaccinated and Unvaccinated Healthcare Workers

| Occupation Code | Frequency | Percent | Cumulative Sum |
|---|---|---|---|
| *Panel A: Vaccinated persons* | | | |
| 221 | 9821 | 10.30 | 10.30 |
| 222 | 1685 | 1.77 | 12.07 |
| 226 | 876 | 0.92 | 12.99 |
| 322 | 29860 | 31.31 | 44.30 |
| 532 | 53113 | 55.70 | 100.00 |
| *Panel B: Unvaccinated persons* | | | |
| 221 | 8293 | 4.27 | 4.27 |
| 222 | 2338 | 1.20 | 5.47 |
| 226 | 3630 | 1.87 | 7.34 |
| 322 | 41385 | 21.30 | 28.64 |
| 532 | 138648 | 71.36 | 100.00 |

*Notes:* This table presents the number of vaccinated and unvaccinated healthcare workers in each occupation. Table A1 provides description for each occupation code. The occupation codes are based on the classification of occupations (TK10) of Statistics Finland.

### D Estimation Method

Vaccine effectiveness was reported as a relative risk reduction (RRR), a widely used metric for assessing vaccine effectiveness in epidemiology. The RRR measures how much the vaccination reduced the cumulative risk of SARS-CoV-2 infection in the treatment group relative to the control group, who did not receive a vaccination. We separately investigated healthcare workers and their unvaccinated family members (spouses and children).

We estimated the following log-binomial model separately for each time-to-event week $l$, defined as the number of periods in calendar week $t$ from obtaining the first dose of vaccination:

$$\ln P_{it} = \alpha_l + \beta_l T_i + X_{it}'\delta + \lambda_{tl} + e_{it} \tag{1}$$

where $P_{it} = P(y_{it} = 1)$ is the probability of the cumulative SARS-CoV-2 infection of individual or household (in the analysis for children) $i$ recorded in calendar week $t$ and thereafter ($y_{it} = 1$). When analysing the indirect effects of vaccination on the children, the outcome variable is coded as one if any child of the healthcare worker living in the same household has SARS-CoV-2 infection and zero otherwise. Moreover, $T_i$ is an indicator variable for treatment group status (vaccinated healthcare worker, spouse or parent), and $\lambda_{tl}$ contains week fixed effects that capture the state of the epidemic in each time-to-event week $l$, and $X_{it}$ contains controls at the individual-level (or for the oldest child): age, age squared and sex. The $\alpha_l$ refers to the log risk in the control group and $\beta_l$ refers to differences in the log risks between the treatment and control group in time-to-event week $l$. The key advantage of a log-binomial model is that it allows easy access to an estimate of the RRR: the RRR is simply $1 - \exp(\beta_l)$. The corresponding 95% confidence interval estimates were calculated using the standard errors of the $\beta_l$ coefficients.

As the individuals in the control group are unvaccinated, we randomly drew the starting date of the follow-up period from the observation period of vaccinations (December 27, 2020 - March 18, 2021) for these unvaccinated individuals. We used this approach in the main specifications to ensure that the vaccine effectiveness estimates are not biased by the changing nature of the epidemic between the follow-up of the treatment and control groups. We also conducted a sensitivity check, where we followed individuals in the control group from the start of the mass vaccination program, December 27, 2020 (Figure A6, Section E).

### E Supplementary Results

The recommended interval between the first and second dose of the mRNA vaccines was initially three weeks but changed to 12 weeks in February 2021. Figure A1 shows the average rate of receiving the second dose after the first dose in our sample. In the first two weeks after the first dose, the share of fully vaccinated individuals was zero, and in week three, it was approximately 35%. In weeks 4-10, over 40% of vaccinated healthcare workers had obtained their second dose.

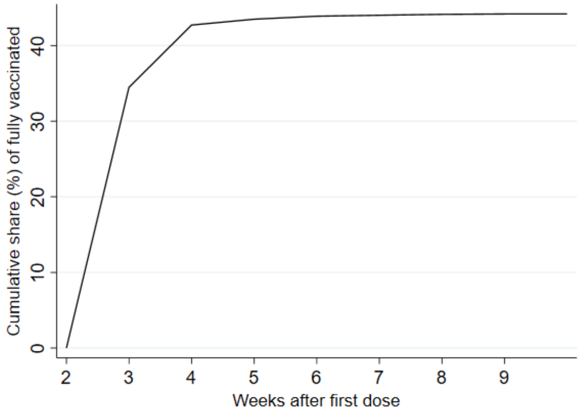


Figure A1: The share of individuals with a second dose in our sample

*Notes:* This figure plots the cumulative share of individuals with a second dose in our sample by week after obtaining the first dose. We calculated the shares by dividing the cumulative number of individuals with a second dose by the number of individuals with at least one dose.

To study the sensitivity of our results for model specification, we estimated vaccine effectiveness using a linear probability model (OLS). The model is similar to that in equation (1), but the outcome is simply the indicator of SARS-CoV-2 infection, $y_{it}$. To calculate the RRR in this case, we divided the $\beta_l$ coefficient by the average probability of $y_{it}$ in the control group in time-to-event week $l$. The 95% confidence intervals were calculated using the standard errors of the $\beta_l$ coefficients, clustered at the individual-level. The results from the linear model for healthcare workers, their unvaccinated spouses and children are similar to the main findings, as shown in Panels A and B of Figures A2 and A3.

Moreover, we estimated linear probability models (so-called difference-in-differences specification) that additionally accounted for the possible pre-existing differences in the outcomes between the treatment and control groups when estimating the treatment effects. Again, we obtain similar results on the vaccine effectiveness for healthcare workers (Figure A3, Panel A), their unvaccinated spouses (Figure A3, Panel B) and children (Figure A5).

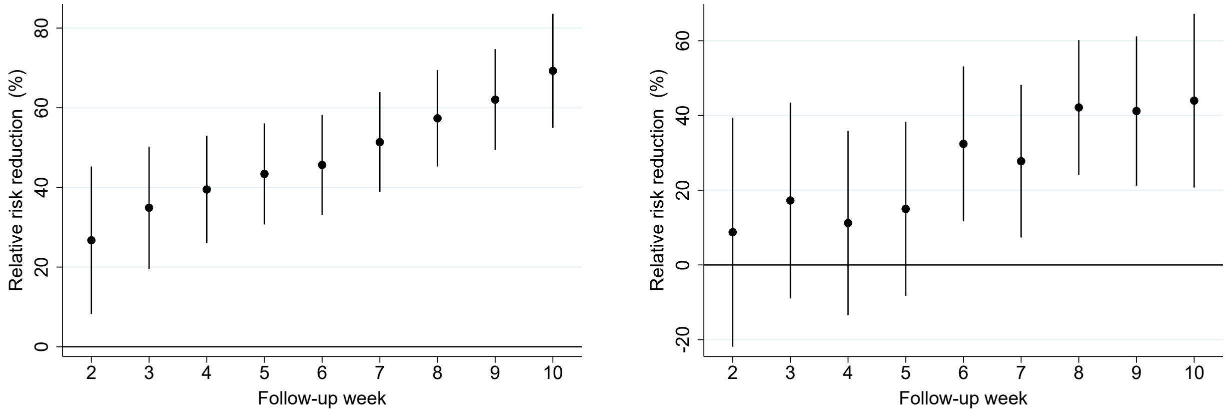


(a) Vaccine effectiveness on vaccinated individuals

(b) Vaccine effectiveness on unvaccinated spouse

Figure A2: Vaccine effectiveness on vaccinated and unvaccinated spouses living in the same household. Coefficients are estimated using the linear probability model and random assignment of the follow-up period in the control group.

*Notes:* This figure plots the vaccine effectiveness estimates by week after the first dose of mRNA-based vaccination. The dependent variable is a polymerase chain reaction (PCR)-confirmed SARS-CoV-2 infection as recorded in the Finnish National Infectious Diseases Register. **Panel A** shows our direct effectiveness estimates (relative risk reduction) for vaccinated individuals compared to the control group, which is constructed by randomly assigning the beginning of the follow-up period for non-vaccinated individuals in the control group. **Panel B** shows the effectiveness estimates (relative risk reduction) for the non-vaccinated spouses (including cohabiting partners) of vaccinated individuals who lived in the same household as the vaccinated person as of December 31, 2019. See Supplementary Appendix for details on sample restrictions. Coefficients in both panels are estimated using individual-date data collapsed to individual-week data. To construct the relative risk reduction estimates, we first regress the PCR-confirmed SARS-CoV-2 infection on a vaccination status indicator variable (vaccinated vs. unvaccinated) by the event week (follow-up week after vaccine administration). We then divide the regression coefficients by control group mean value. The solid black dots show relative risk reduction by week after receiving the first dose of mRNA-based vaccination. The error bars denote the 95% confidence intervals clustered at the individual level using endpoint transformation of originally estimated confidence intervals.

Finally, we explore the robustness of our findings by estimating alternative log-binomial models, where we followed all individuals in the control group from the beginning of the mass vaccination program (December 27, 2020). The disadvantage of this specification is that the effectiveness estimates may reflect changes in the state of the epidemic from the beginning of the vaccination program, rather than the true effectiveness

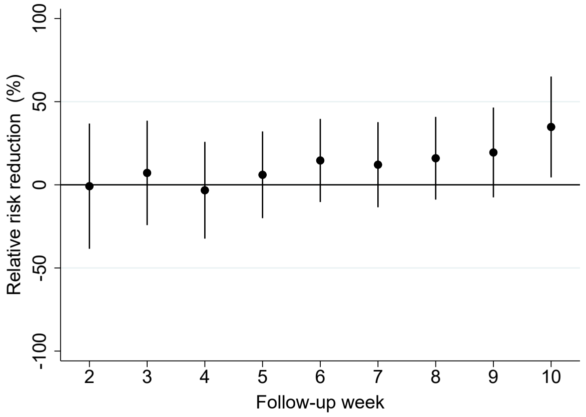


(a) Vaccine effectiveness on unvaccinated household members aged from 3 to 18 years

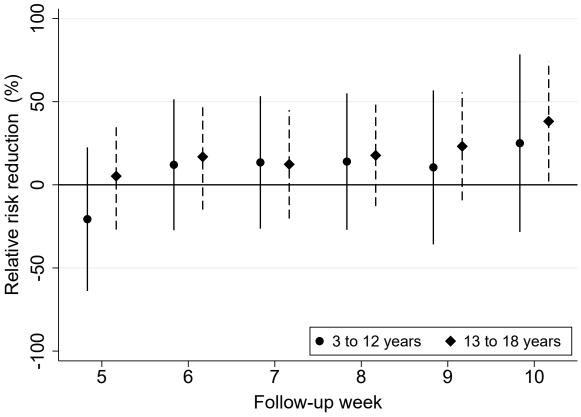


(b) Vaccine effectiveness on unvaccinated household members by age group

Figure A3: Vaccine effectiveness on unvaccinated children and adolescents. Coefficients are estimated using the linear probability model and random assignment of the follow-up period in the control group.

*Notes:* This figure plots the vaccine effectiveness estimates by week after the first dose of mRNA-based vaccination. The dependent variable is a polymerase chain reaction (PCR)-confirmed SARS-CoV-2 infection as recorded in the Finnish National Infectious Diseases Register. **Panel A** shows effectiveness estimates (relative risk reduction) for unvaccinated household members aged from 3 to 18 years who lived in the same household with the vaccinated person as of December 31, 2019. **Panel B** shows effectiveness estimates (relative risk reduction) separately for unvaccinated children aged from 3 to 12 years and adolescents aged from 13 to 18 years who lived in the same household with the vaccinated person as of December 31, 2019. Panel B stacks the estimates from follow-up weeks 2 to 5 to a single estimate as there are too few polymerase chain reaction (PCR)-confirmed SARS-CoV-2 infections among children aged from 3 to 12 years to make any inference about the short-term effectiveness of mRNA-based vaccination in this age group. See Online Appendix, Section C, for details on sample restrictions. Coefficients in both panels are estimated using individual-date data collapsed to individual-week data. To construct the relative risk reduction estimates, we first regress the PCR-confirmed SARS-CoV-2 infection on a vaccination status indicator variable (parent vaccinated vs. parents unvaccinated) by the event week (follow-up week after vaccine administration). We then divide the regression coefficients by control group mean value. The solid black dots show relative risk reduction by week after parent receiving the first dose of mRNA-based vaccination. The error bars denote the 95% confidence intervals clustered at the individual level using endpoint transformation of originally estimated confidence intervals.

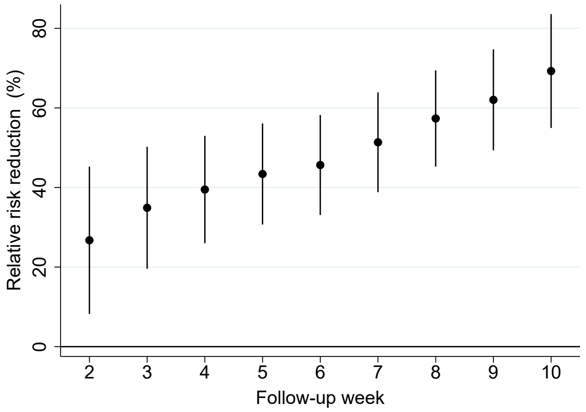


(a) Vaccine effectiveness on vaccinated individuals

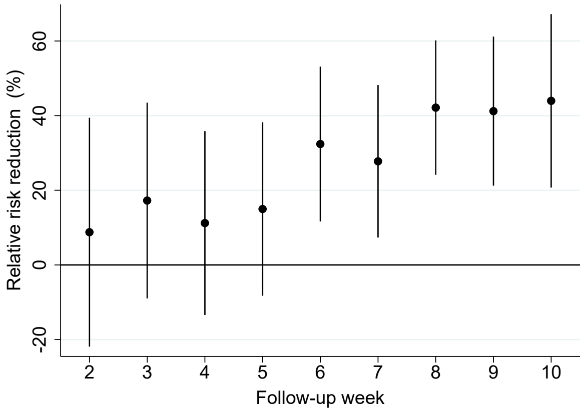


(b) Vaccine effectiveness on unvaccinated spouse

Figure A4: Vaccine effectiveness on vaccinated individuals and unvaccinated spouses living in the same household. Coefficients are estimated using the differences-in-differences model and random assignment of the follow-up period in the control group.

*Notes:* This figure plots the vaccine effectiveness estimates by week after receiving the first dose of mRNA-based vaccination. The dependent variable is a polymerase chain reaction (PCR)-confirmed SARS-CoV-2 infection as recorded in the Finnish National Infectious Diseases Register. **Panel A** shows the effectiveness estimates (relative risk reduction) for vaccinated individuals compared to the control group, which is constructed by randomly assigning the beginning of the follow-up period for unvaccinated individuals in the control group. **Panel B** shows the effectiveness estimates (relative risk reduction) for the unvaccinated spouses (including cohabiting partners) of vaccinated individuals who lived in the same household as the vaccinated person as of December 31, 2019. See Supplementary Appendix for details on sample restrictions. Coefficients in both panels are estimated using individual-date data collapsed to individual-week data. To construct the relative risk reduction estimates, we first regress the PCR-confirmed SARS-CoV-2 infection on the interaction of event week (follow-up week after vaccine administration) and vaccination status (vaccinated vs. unvaccinated). We then divide the regression coefficients by control group mean value. The solid black dots show relative risk reduction by week after receiving the first dose of mRNA-based vaccination. The error bars denote the 95% confidence intervals clustered at the individual-level using endpoint transformation of originally estimated confidence intervals.

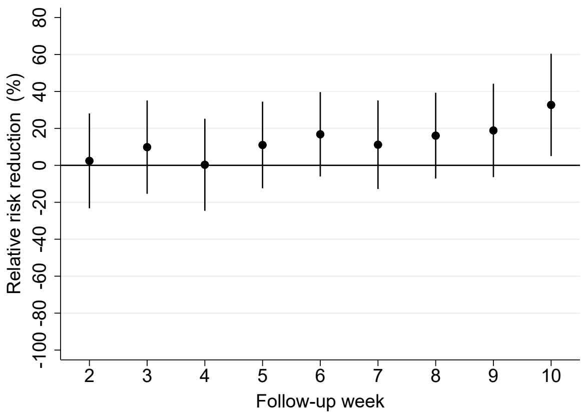


(a) Vaccine effectiveness on unvaccinated household members aged from 3 to 18 years

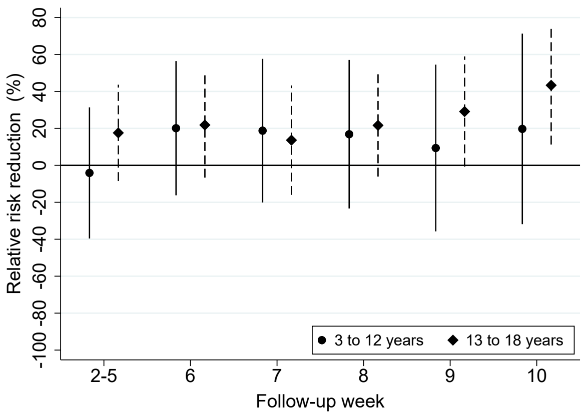


(b) Vaccine effectiveness on unvaccinated household members by age groups

Figure A5: Vaccine effectiveness on unvaccinated children and adolescents. Coefficients are estimated using the differences-in-differences model and random assignment of the follow-up period in the control group.

*Notes:* This figure plots the vaccine effectiveness estimates by week after the first dose of mRNA-based vaccination. The dependent variable is a polymerase chain reaction (PCR)-confirmed SARS-CoV-2 infection as recorded in the Finnish National Infectious Diseases Register. **Panel A** shows effectiveness estimates (relative risk reduction) for unvaccinated household members aged from 3 to 18 years who lived in the same household with the vaccinated person as of December 31, 2019. **Panel B** shows effectiveness estimates (relative risk reduction) separately for unvaccinated children aged from 3 to 12 years and adolescents aged from 13 to 18 years who lived in the same household with the vaccinated person as of December 31, 2019. Panel B stacks the estimates from follow-up weeks 2 to 5 to a single estimate as there are too few polymerase chain reaction (PCR)-confirmed SARS-CoV-2 infections among children aged from 3 to 12 years to make any inference about the short-term effectiveness of mRNA-based vaccination in this age group. See Online Appendix, Section C, for details on sample restrictions. Coefficients in both panels are estimated using individual-date data collapsed to individual-week data. To construct the relative risk reduction estimates, we first regress the PCR-confirmed SARS-CoV-2 infection on a vaccination status indicator variable (parent vaccinated vs. parents unvaccinated) by the event week (follow-up week after vaccine administration). We then divide the regression coefficients by control group mean value. The solid black dots show relative risk reduction by week after parent receiving the first dose of mRNA-based vaccination. The error bars denote the 95% confidence intervals clustered at the individual level using endpoint transformation of originally estimated confidence intervals.

of vaccination. The results for healthcare workers are qualitatively consistent with the results from our main specification using a randomly assigned follow-up period for the control group (Figure A6, Panel A). For the spouses and children, the trend in vaccine effectiveness is similar to that observed in the main specification, but the point estimates suggest negative effectiveness in the first follow-up weeks with notable statistical imprecision (Figures A6, Panel B and Figure A7). The children's negative effectiveness estimates seem to be driven by the children aged 3-12 years old (Figure A7, Panel A vs Panel B).

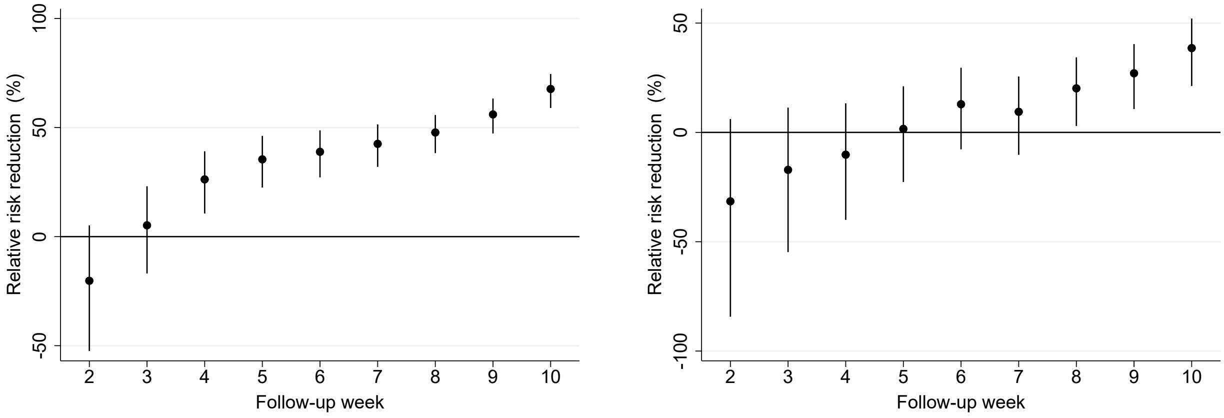


(a) Vaccine effectiveness on vaccinated individuals

(b) Vaccine effectiveness on unvaccinated spouse

Figure A6: Vaccine effectiveness on vaccinated individuals and unvaccinated spouses living in the same household. Coefficients are estimated using a log-binomial model and following all individuals in the control group from the beginning of the mass vaccination program (December 27, 2020)

*Notes:* This figure plots the vaccine effectiveness estimates by week after the first dose of mRNA-based vaccination. The dependent variable is a polymerase chain reaction (PCR)-confirmed SARS-CoV-2 infection as recorded in the Finnish National Infectious Diseases Register. **Panel A** shows our direct effectiveness estimates (relative risk reduction) for vaccinated individuals compared to the control group, which is constructed by following all individuals in the control group from the beginning of the mass vaccination program (December 27, 2020). **Panel B** shows the effectiveness estimates (relative risk reduction) for the non-vaccinated spouses (including cohabiting partners) of vaccinated individuals who lived in the same household as the vaccinated person as of December 31, 2019. See Supplementary Appendix for details on sample restrictions. Coefficients in both panels are estimated using individual-date data collapsed to individual-week data. To construct the relative risk reduction (RRR) estimates, we first run a log-binomial regression model using a vaccination status indicator variable (vaccinated vs. unvaccinated) by the event week (follow-up week after vaccine administration) and transform the obtained estimates into risk ratio coefficients. We then compute the relative risk reduction using the formula RRR = 1 - risk ratio. The solid black dots show relative risk reduction by week after receiving the first dose of mRNA-based vaccination. The error bars denote the 95% confidence intervals clustered at the individual level using endpoint transformation of originally estimated confidence intervals.

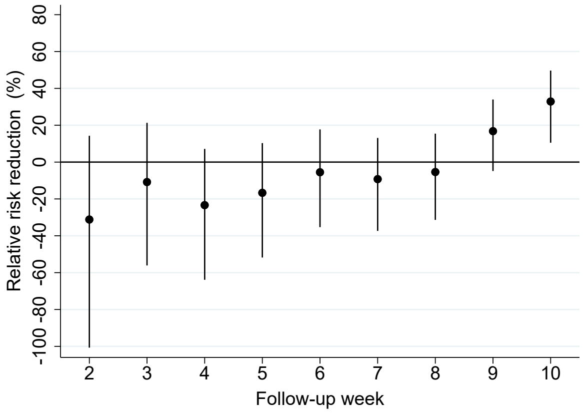


(a) Vaccine effectiveness on unvaccinated household members aged from 3 to 18 years

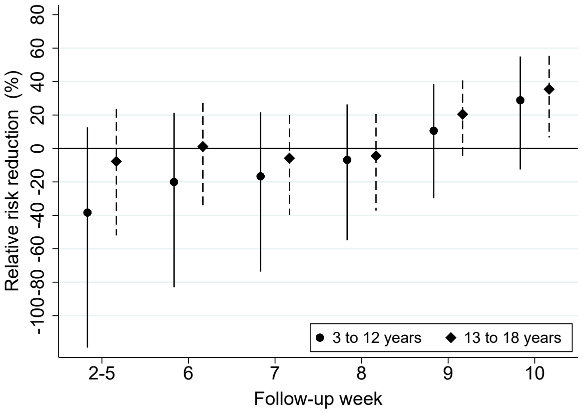


(b) Vaccine effectiveness on unvaccinated household members by age groups

Figure A7: Vaccine effectiveness on unvaccinated children and adolescents. Coefficients are estimated using a log-binomial model and following all individuals in the control group from the beginning of the mass vaccination program (December 27, 2020)

*Notes:* This figure plots the vaccine effectiveness estimates by week after the first dose of mRNA-based vaccination. The dependent variable is a polymerase chain reaction (PCR)-confirmed SARS-CoV-2 infection as recorded in the Finnish National Infectious Diseases Register. **Panel A** shows effectiveness estimates (relative risk reduction) for unvaccinated household members aged from 3 to 18 years who lived in the same household with the vaccinated person as of December 31, 2019. **Panel B** shows effectiveness estimates (relative risk reduction) separately for unvaccinated children aged from 3 to 12 years and adolescents aged from 13 to 18 years who lived in the same household with the vaccinated person as of December 31, 2019. Panel B stacks the estimates from follow-up weeks 2 to 5 to a single estimate as there are too few polymerase chain reaction (PCR)-confirmed SARS-CoV-2 infections among children aged from 3 to 12 years to make any inference about the short-term effectiveness of mRNA-based vaccination in this age group. See Online Appendix, Section C, for details on sample restrictions. Coefficients in both panels are estimated using individual-date data collapsed to individual-week data. To construct the relative risk reduction (RRR) estimates, we first run a log-binomial regression model using a vaccination status indicator variable (parent vaccinated vs. parents unvaccinated) by the event week (follow-up week after vaccine administration) and transform the obtained estimates into risk ratio coefficients. We then compute the relative risk reduction using the formula RRR = 1 - risk ratio. The solid black dots show relative risk reduction by week after parent receiving the first dose of mRNA-based vaccination. The error bars denote the 95% confidence intervals clustered at the individual level using endpoint transformation of originally estimated confidence intervals.
